# An integrative multi-omics analysis of 16 autoimmune diseases and cancer outcomes highlights immune-cell regulatory mechanisms and shared genetic architecture

**DOI:** 10.1101/2020.11.27.20235663

**Authors:** C Prince, R. E Mitchell, T. G. Richardson

## Abstract

**Background:** Developing functional understanding into the causal molecular drivers of immunological disease is a critical challenge in genomic medicine. Here we systematically apply Mendelian randomization (MR), genetic colocalization, immune cell-type enrichment and phenome-wide association methods to investigate the effect of genetically predicted gene expression on 12 autoimmune and 4 cancer outcomes.

**Results:** Using whole blood derived estimates for regulatory variants from the eQTLGen consortium (n=31,684) we constructed genetic risk scores (r2<0.1) for 10,104 genes. Applying the inverse-variance weighted Mendelian randomization method transcriptome-wide whilst accounting for linkage disequilibrium structure identified 773 unique genes with evidence of a genetically predicted effect on at least one disease outcome (P<4.81 × 10^−5^). We next undertook genetic colocalization to investigate whether these effects may be confined to specific cell-types using gene expression data derived from 18 types of immune cells. This highlighted many cell-type dependent effects, such as *PRKCQ* expression and asthma risk (posterior probability of association (PPA)=0.998), which was T-cell specific, as well as *TPM3* expression and prostate cancer risk (PPA=0.821), which was restricted to monocytes. Phenome-wide analyses on 320 complex traits allowed us to explore the shared genetic architecture and prioritize key drivers of disease risk, such as *CASP10* which provided evidence of an effect on 7 cancer-related outcomes. Similarly, these evaluations of pervasive pleiotropy may be valuable for evaluations of therapeutic targets to help identify potential adverse effects.

**Conclusions:** Our atlas of results can be used to characterize known and novel loci in autoimmune disease and cancer susceptibility, both in terms of developing insight into cell-type dependent effects as well as dissecting shared genetic architecture and disease pathways. As exemplar, we have highlighted several key findings in this study, although similar evaluations can be conducted interactively at http://mrcieu.mrsoftware.org/immuno_MR/.

## Background

Deciphering the genetic architecture of complex traits and disease is a critical challenge for genomic medicine. The widespread application of genome-wide association studies (GWAS) has had profound success in detecting robust associations between genetic variants and complex disease outcomes, including those with a large immunological basis, such as rheumatoid arthritis, inflammatory bowel disease and different types of cancer. (1) There is now extensive interest in the field of genetic epidemiology in integrating findings from GWAS with regulatory molecular datasets. In doing so, studies aim to bring to light the underlying functional and biological mechanisms which may help us understand GWAS signals and translate findings for disease prevention purposes.

A challenge encountered by these endeavours is obtaining molecular trait datasets derived from tissues and cell-types relevant to the disease being studied in sufficient samples. A recent review highlights this by comparing the differences between affected and unaffected tissues for heritable traits and diseases, (2) demonstrating that molecular traits such as gene expression can have exclusive or preferential expression in disease-relevant tissues types. This diminishes the utility of whole blood-derived datasets, which to date typically have by far the largest sample sizes on molecular traits due to their non-invasive accessibility. Notable exceptions to this are disease outcomes with a large immune basis, given that whole blood is responsible for carrying innate and adaptive immune cells through the body from the lymphatic system to the site of injury or infection. (3) As such, initial analyses of transcriptomic datasets derived from whole blood provides optimal statistical power to detect association signals for immune system related disease (4, 5), which can then be dissected and characterized in detail using cell-type specific data. This is particularly important when investigating autoimmune diseases and cancers to develop mechanistic insight into the cell-types which play a role in the causal pathway for these outcomes. (6)

Furthermore, autoimmune diseases and types of cancer are both disorders that affect the immune system. This emphasises the importance of evaluating the shared genetic architecture of these outcomes to highlight the predominating pathways which contribute to risk of multiple types of disease. For example, previous work in this area has identified evidence of shared architecture between Crohn’s disease and multiple sclerosis, (7) across several paediatric autoimmune diseases, (8) and also across cancer outcomes due to immune-related mechanisms. (9) Moreover, genetic evidence of horizontal pleiotropy at loci which encode a therapeutic target may be informative in terms of flagging potential adverse effects unrelated to autoimmune and cancer outcomes, which is particularly attractive given the increasing interest in using human genetics to help validate drug targets. (10)

In this study, we have applied the principles of Mendelian randomization (MR) to evaluate genetically predicted effects of gene expression derived from whole blood on 12 autoimmune and 4 cancer outcomes. MR is a form instrumental variable analysis which uses genetic variants to infer causal relationships between exposures and disease outcomes, which are more robust to confounding and reverse causation given that they are inherited at birth. (11, 12) As MR studies of molecular traits have typically been limited to single instrument analyses in the past, here we applied a more novel approach using regulatory variants as genetic instruments (r^2^ <0.1) from the eQTLGen consortium (n=31,684) whilst accounting for their linkage disequilibrium (LD) structure in the analysis framework. We then explored whether putative genetics effects identified using whole blood may be cell-type specific using expression data from 18 different immune-cell regulatory datasets from the BLUEPRINT consortium and DICE database using genetic colocalization. (13, 14) Finally, we undertook a phenome-wide association study (PheWAS) of genes highlighted by these analyses to assess their shared architecture and pathways using data on a total of 320 curated complex traits and outcomes. A diagram of this analysis pipeline can be found in Figure 1.

**Figure 1.**
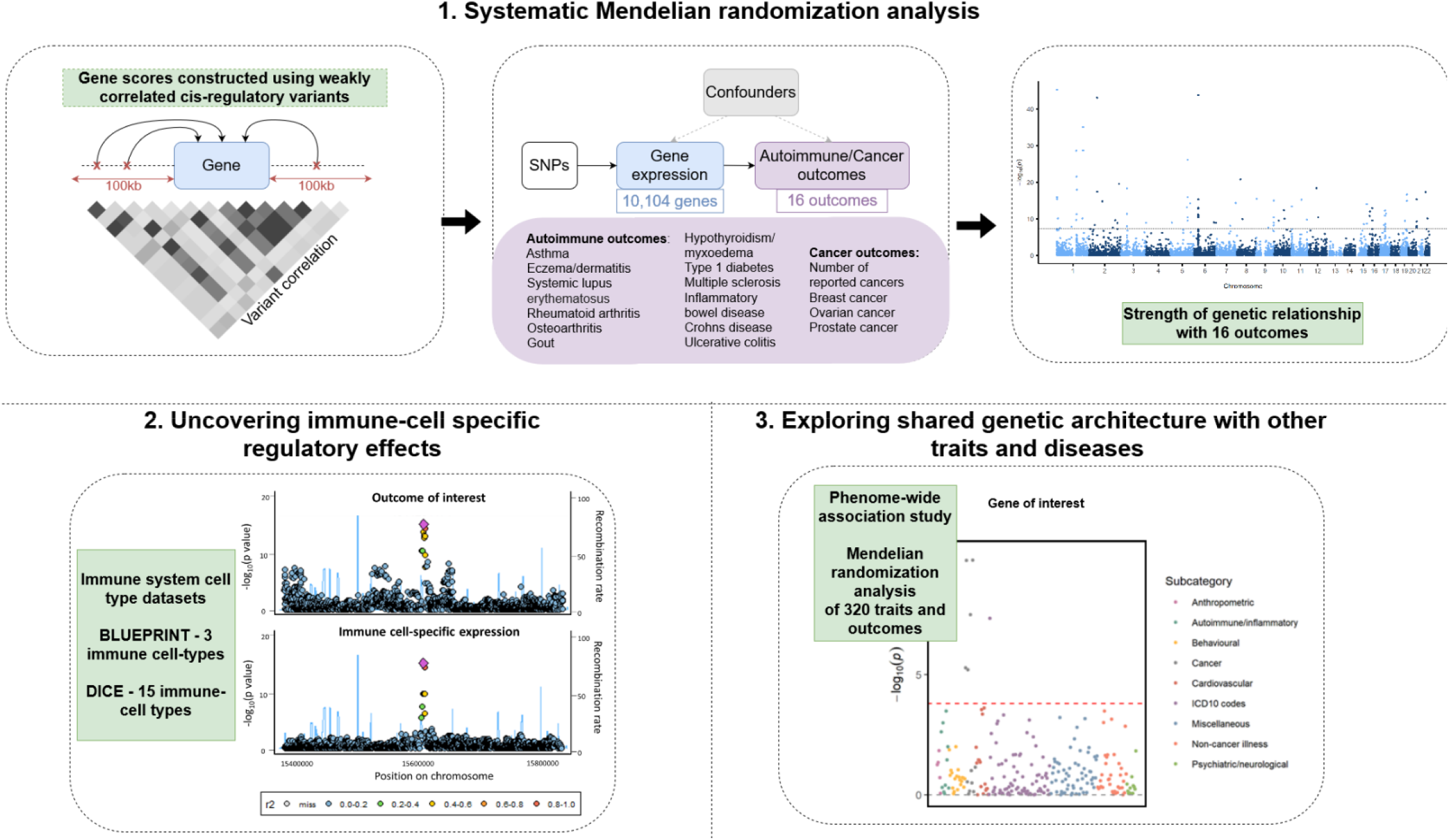
Outline of study workflow. 1. First, a MR approach using weakly correlated instruments was applied to investigate the genetically predicted effects of 10,104 genes on 16 outcomes. 2. A genetic colocalization method was performed to investigated whether gene expression in specific cell-types at identified loci shares a common causal variant with autoimmune or cancer disease risk using BLUEPRINT and DICE immune cell datasets. 3. Finally a similar MR approached was applied to a curated set of 320 health outcomes, to assess shared genetic architecture and where identified genes were likely to be involved in multiple biological pathways.

## Results

### An integrative Mendelian randomization analysis of 16 autoimmune diseases and cancer outcomes using multiple cis-regulatory instruments

Using effect estimates derived from whole blood from the eQTLGen consortium (n=31,684), we undertook LD clumping to construct genetic scores using weakly independent cis-regulatory variants based on r^2^ <0.1. LD calculations were based on a reference panel of 10,000 unrelated UK Biobank participants of European descent. (15-17) We then applied MR using these gene-based scores to systematically investigate the genetically predicted effects of each gene in turn on risk of 16 disease outcomes (12 autoimmune diseases and 4 cancer-related outcomes) using summary data from GWAS (**Supplementary Table 1**) (18-26). This was undertaken using the inverse variance weighted (IVW) MR method which estimates genetically predicted effects whilst accounting for LD structure between all genetic instruments used in the score (16, 27). After LD clumping, there were 10,104 genes which had at least 2 weakly independent cis-QTLs that were eligible for analysis using the IVW method. (**Supplementary Table 2**)

In total, we identified 827 genes which provided evidence of a genetically predicted effect on at least 1 outcome after accounting for multiple testing using the Bonferroni corrected threshold for each outcome separately (ranging from P=4.09×10^−6^ to 4.81×10^−5^) (**Supplementary Table 3-4**). Of these, 773 were located outside the human leukocyte antigen (HLA) region of the genome and were carried forward for subsequent analyses due to the extensive LD structure at HLA which can results in false positive findings when using techniques such as genetic colocalization. **Figure 2** illustrates various exemplar signals identified for 4 outcomes: asthma, hypothyroidism, breast cancer and inflammatory bowel disease. Full results for all 16 outcomes can be investigated using the interactive web browser available at http://mrcieu.mrsoftware.org/immuno_MR/.

**Figure 2.**
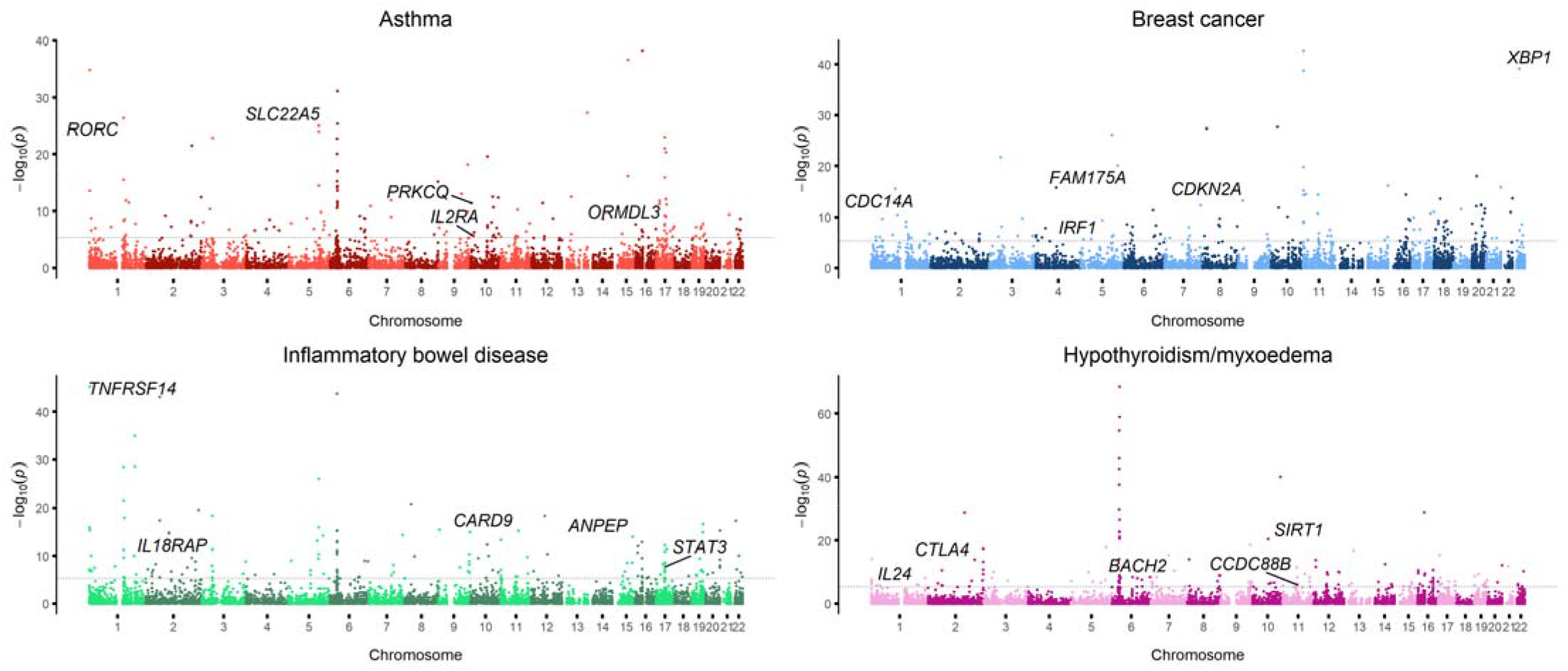
Manhattan plots illustrating the association between gene expression and 4 outcomes: asthma, hypothyroidism/myxoedema, breast cancer and inflammatory bowel disease. Bonferroni corrected threshold, indicated by the dotted line, was used to identified strong associations.

Amongst these results were genetically predicted effects at various well-established loci known to confer risk of autoimmune disease, including *CARD9* and *STAT3* (P= 1.03×10^−15^ and 1.77 x10^−8^ respectively with inflammatory bowel disease), (28) *ORMDL3* associated with asthma (P= 4.82 x10^−10^), (29) and cytokines such as interleukin-24 (*IL24*) and interleukin-2 receptor alpha chain (*IL2RA*) which were associated with systemic lupus erythematosus and asthma respectively (P= 1.28 x10^−6^ and 1.34 x10^−6^). (30, 31) A number of novel or emerging loci were also identified for autoimmune disease outcomes, such as *RORC*, a transcription factor predominantly expressed in T helper 17 cells (32) which was most strongly associated with asthma risk (P=4.13 x10^−27^), as well as *CCDC88B* (P= 1.07 x10^−6^ and 1.15 x10^−5^ with hypothyroidism/myxoedema and multiple sclerosis respectively). These findings were additionally supported by evidence from our leave-one-out sensitivity analysis to highlight signals which were not dependent on single cis-instruments. (**Supplementary Table 5**)

Similarly, there were various signals at known cancer loci, such as *CASP10* (P= 1.82 x10^−17^ for prostate cancer), (*33*) and *FAM175A* (P= 1.56 x10^−16^ for breast cancer), (*34*) as well as genes that have been identified in relation to a number of cancers including *CDKN2A* (P= 5.08 x10^−14^ with breast cancer), *IRF1* (P= 7.40×10^−7^ with breast cancer) and *IGF2* (P= 1.67 x10^−6^ with prostate cancer). (35) There were also loci highlighted by our analyses on cancer outcomes with limited previous evidence of an association with cancer outcomes based on the current literature and therefore may be more novel, such as *PSMD8* (P= 5.60 x10^−9^ with prostate cancer) and *TTC16* (P= 1.10 x10^−9^ with prostate cancer). *PSMD8* is a proteasome subunit has been identified in the regulation of the cell growth and differentiation and apoptosis. Targeting the proteasome has recently been postulated as a potential cancer therapy. (36, 37)

### Identifying immune-cell specific effects at autoimmune and cancer associated loci

We performed genetic colocalization at each of the 773 non-HLA loci identified in the previous analysis using 15 immune-cell datasets from the DICE database and 3 immune cell-type datasets from the BLUEPRINT consortium. This was to evaluate whether gene expression in specific cell-types at these loci share a common causal variant with autoimmune or cancer disease risk. This was performed using the ‘coloc’ R package, where a posterior probability of association (PPA) threshold of >0.8 was used to indicate evidence of colocalization. (38)

In total, 538 genetic effects colocalised across the cancer and autoimmune disease outcomes with immune-cell type expression which may provide mechanistic insight into the disease pathogenesis at associated loci. For example, we identified strong evidence of colocalization between *PRKCQ* expression in T cells and asthma risk (PPA=0.998), whereas there was very weak evidence of colocalization when analysing any of the other immune cell-types (**Figure 3a, Supplementary Table 7**). *PRKCQ* has been previously implicated in allergic disease risk and is involved in T cell activation. (39) There was also evidence of colocalization between *KSR1* expression and Crohn’s disease in classical monocytes (PPA= 0.998) (**Figure 3b, Supplementary Table 7**), which is known to be an important cell-type in relation to Crohn’s disease. (40) Amongst cancer loci, there was evidence for colocalization between prostate cancer and *C2orf43* in non-classic monocytes (PPA=0.887)) (**Figure 3c, Supplementary Table 7**). *C2orf43* has been found to be expressed in monocytes and the loss of this gene has previously been associated with risk of prostate cancer. (41, 42) All other effects with evidence of genetic colocalization are shown in **Supplementary Table 6** as well as on our web browser where effects across all cell-type can be compared visually.

**Figure 3.**
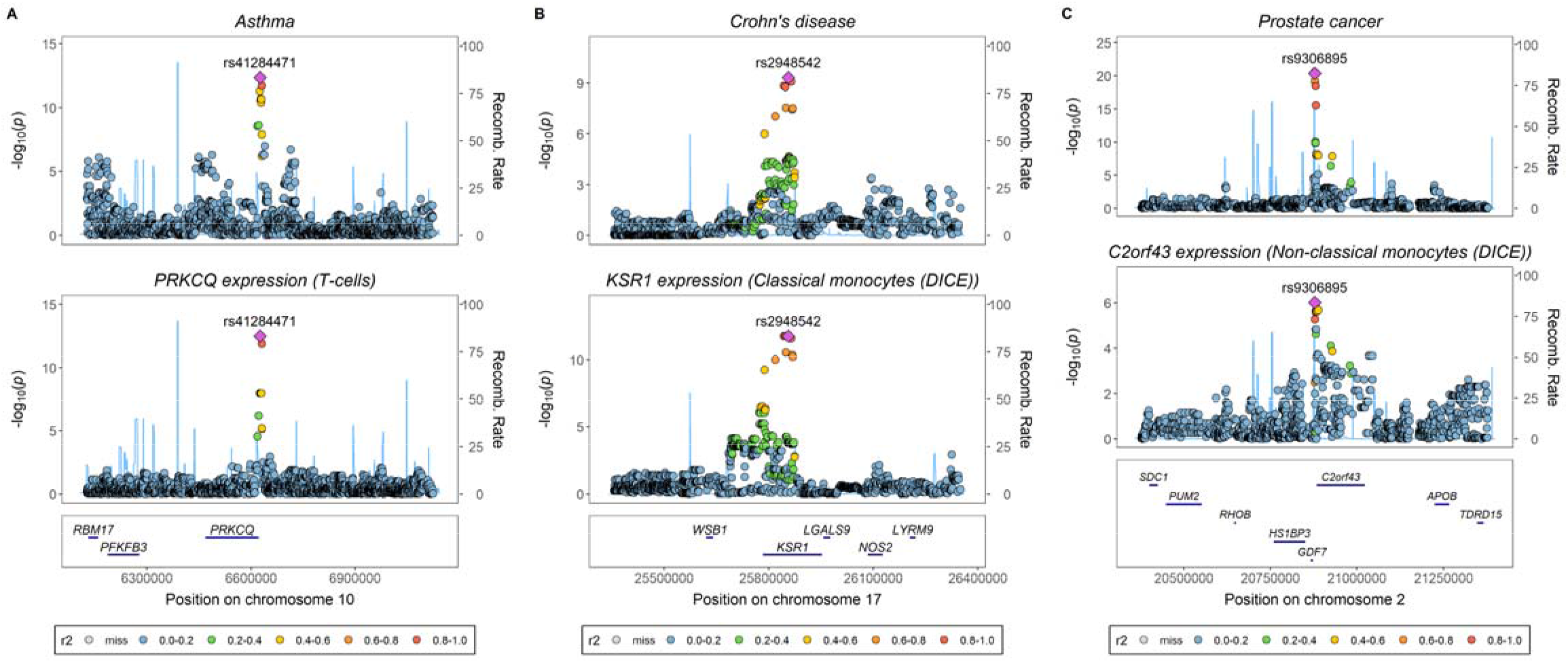
Locuszoom plot illustrating colocalization between A) Asthma and PRKCQ expression in T cells, B) Crohn’s disease and KSR1 expression in classical monocytes and C) Prostate cancer and C2orf43 expression in non-classical monocytes.

### Enrichment of immune-cell types amongst disease-associated loci

We performed enrichment analyses using results from the colocalization analyses to determine whether effects in certain immune-cell types were overrepresented amongst each outcome, and whether certain cell types were key for each outcome. We did not include the number of reported cancers results in the background set as there was no strong evidence of colocalization for any gene using this outcome, which may reflect that associated loci are more likely to be involved in risk factors for cancer rather than being directly involved in cancer pathogenesis themselves.

As illustrated in **Figure 4**, we identified evidence of enrichment for various cell-types amongst rheumatoid arthritis loci and in particular for activated naïve CD8 T cells (P= 1.79 x10^−4^). Increased levels of these cells have been previously observed in the peripheral blood of patients with rheumatoid arthritis. (43) Monocytes were enriched amongst multiple sclerosis loci (P= 6.34 x10^−4^) which have previously been implicated in the pathology of this disease. (44) Regulatory memory T cells were enriched for breast cancer loci (P= 0.018); which have previously been identified as markers of poor prognosis of breast cancer. (45) (**Supplementary Tables 8-9**)

**Figure 4.**
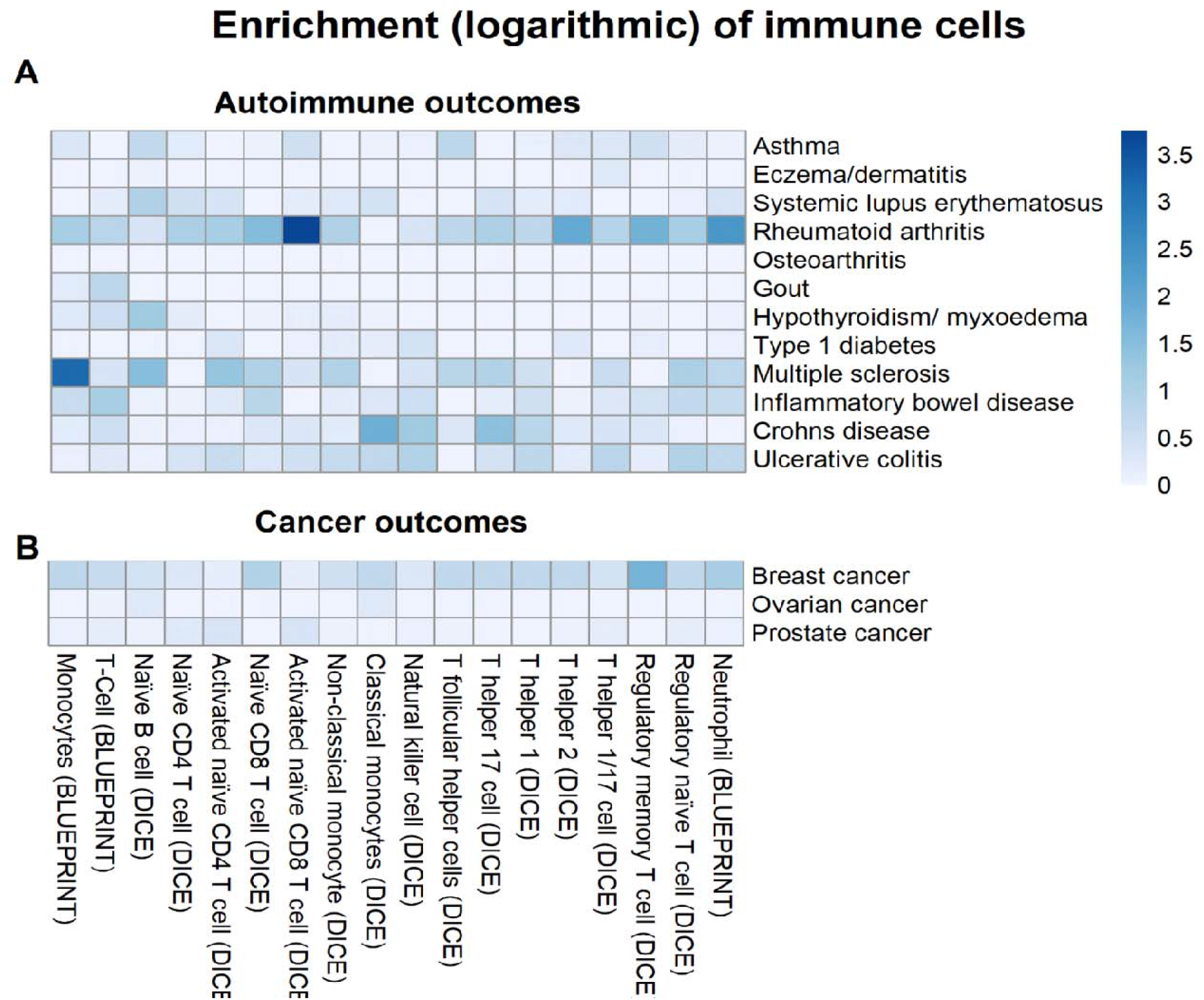
Enrichment plot illustrating for which immune-cell types were effects were overrepresented amongst each outcome.

### Conducting phenome-wide association studies to explore shared genetic architecture and elucidate pleiotropic loci

For the 773 non-HLA genes identified in our primary analysis using whole blood, we repeated analyses using the IVW MR analysis accounting for LD structure but on a set of 320 curated traits and outcomes. These included the initial 16 autoimmune and cancer outcomes as well as 304 traits related to broad range of outcomes from across the complex disease spectrum (**Supplementary Table 10**). This phenome-wide analysis allowed us to highlight loci where there is evidence of shared genetic architecture amongst various autoimmune and cancer outcomes. For instance, *IL24*, which encodes an interleukin cytokine involved in promoting the development and differentiation of T, B, and hematopoietic cells, and plays an essential role in both innate and adaptive immunity, (46) provided evidence of an effect on multiple autoimmune outcomes (**Figure 5a**). Similarly, analyses of *CASP10*, which encodes caspase 10 and is a known cancer susceptibility locus, identified genetically predicted effects on 7 different cancer disease outcomes. (33) (**Figure 5b**) There was also evidence of shared architecture at emerging immune disease loci, such as *CCDC88B*, which has recently been implicated in the pathogenesis of inflammatory bowel disease. (47)

**Figure 5.**
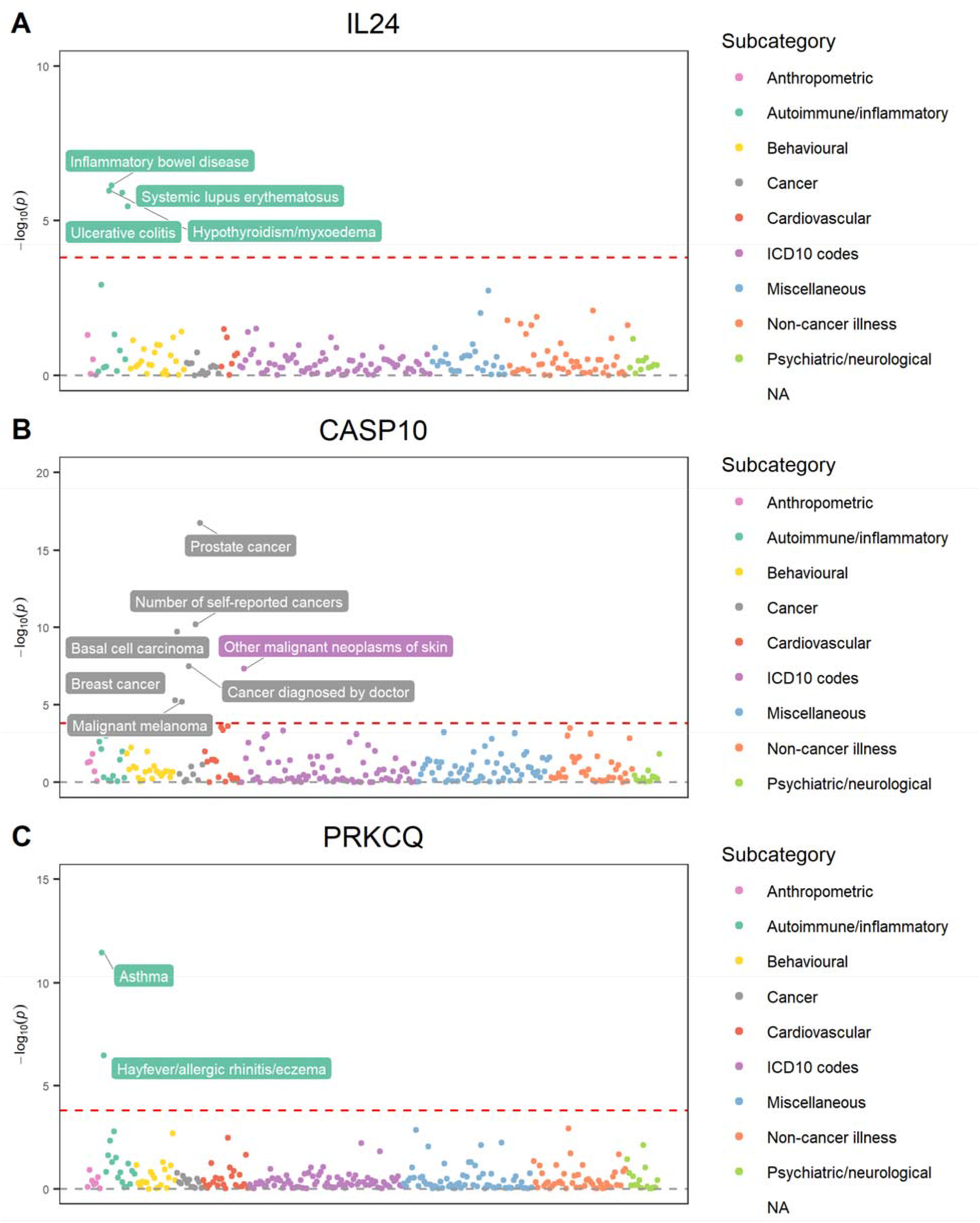
Phenome-wide association study (PheWAS) plots illustrating genetically predicted effects between expression of A) IL24, B) CASP10 and C) PRKCQ, and 320 health outcomes.

Along with evaluations of loci with shared architecture for immune-related outcomes, our atlas of phenome-wide results may be valuable in highlighting genes with more specific effects on disease outcomes. For example, *PRKCQ* was highlighted by our cell-type analysis as having a T-cell specific mediated effect on asthma risk (PPA=0.998), and only provided robust evidence of an effect on allergic disease (P=3.41×10^−7^) along with asthma as discovered in our initial analysis based on the number of tests undertaken (P<1.56×10^−4^=0.05/320 tests) (**Figure 5c**). Similar evaluations of pleiotropy may have translatable benefit for drug target prioritization efforts. For instance, *TPM3* has recently been postulated as a potential therapeutic target for cancer therapy. (48) Although our cell-type analysis detected evidence of a monocyte-specific role of *TPM3* in prostate cancer risk (PPA=0.821), phenome-wide results indicated that it may influence risk of outcomes such as hypertension (P=1.49×10^−7^) and angina (P=2.81×10^−9^) with the opposite direction of effect. These results therefore suggest that loci which exhibit horizontal pleiotropic effects such as *TPM3* should be deprioritised as therapeutic targets due to putative adverse effects. Results depicted in Figure 5 can also be found in **Supplementary Table 11-14**. Interactive phenome-wide results can be visualised using our web browser.

## Discussion

In this study, we have performed a transcriptome-wide Mendelian randomization study to investigate the genetically predicted effects of gene expression on risk of 12 autoimmune diseases and 4 cancer outcomes. The results of this investigation provide a comprehensive atlas of genetic effects which highlight both known and novel susceptibility loci for these outcomes. We conducted in-depth analyses of these loci using genetic colocalization and phenome-wide MR to further characterize their role in disease, both in terms of developing mechanistic insight into cell-type dependent effects as well as elucidating shared biological pathways. As exemplar, we have highlighted several key findings in this manuscript, however all our results can be investigated interactively at http://mrcieu.mrsoftware.org/immuno_MR/. We envisage this atlas of results will benefit future research endeavours concerned with dissecting the molecular drivers of autoimmune disease and cancer outcomes, as well as help guide functional studies to validate and strengthen evidence for loci highlight in our study.

Integrating molecular regulatory signatures derived from whole blood with findings from GWAS has been considered a limitation for the majority of complex disease outcomes studied to date. (49) However, it presents a viable strategy for immune system-related diseases given that whole blood is responsible for carrying innate and adaptive immune cells through the body from the lymphatic system to the site of injury or infection (3). This has allowed us to harness the unparalleled sample size of transcriptome-wide data made available by the eQTLGen consortium. As a consequence, we were able to instrument genes using multiple regulatory variants and address another conventional limitation of previous studies in the paradigm which have typically been confined to single-SNP MR analyses. (50) Overall, 773 genes passed our Bonferroni corrected threshold excluding those in the HLA region of the genome.

Amongst these findings are many previously reported autoimmune disease and cancer genes. For example, *CARD9*, identified here in relation to Crohn’s disease and ulcerative colitis, has previously been reported to confer risk of both these forms of inflammatory bowel disease, (51) and is known to be involved in innate immunity and inflammation, as well as being specifically expressed in myeloid cells. (52) Likewise, *STAT3* has been identified in relation to inflammatory bowel diseases and type 1 diabetes and is thought to be involved in autoimmunity both due to its role as a mediator on the IL-6 signalling pathway and as a transcription factor in the differentiation of Th1 cells. (53, 54) Amongst established cancer loci was *CTBP1* which we identified evidence as having a genetically predicted effect on breast and prostate cancer risk. *CTBP1* is an oncogenic transcriptional co-regulator which has been shown to be overexpressed in a number of cancers. It functions by regulating the expression of tumour suppressers and oncogenic factors, which has led to its identification as a potential therapeutic target. (55)

There were also less well-established loci identified in the MR analysis. *RORC* was identified in relation to inflammatory bowel diseases, osteoarthritis, eczema and asthma. It is a transcription factor of *IL-17* expression and Th17 cells, which are key in the immune system and has been suggested as a potential target for autoimmune diseases. (56) *IL-17* is a pro inflammatory cytokine which recruits immune cells to the site of inflammation and it’s overproduction has been reported to lead to inflammation leading to autoimmune conditions. (57, 58) *CCDC88B* was identified in relation to hypothyroidism/myxoedema and multiple sclerosis, and has previously been shown to be highly expression in immune cells. It has been identified as an important regulator of T cell function and previously implicated to play a role in inflammation pathways. (59)

Applying genetic colocalization revealed many cell-type dependent effects in our study, which may help develop understanding into the pathways and mechanisms behind these disease outcomes. For example, there was strong evidence of colocalization between *CARD9* expression and inflammatory bowel diseases in monocytes (PPA= 0.986) and neutrophils (PPA= 0.986). The effect of *KSR1* expression on risk of Crohn’s disease colocalized with data from monocytes (PPA= 0.998). *KSR1*, which encodes a kinase suppressor of Ras 1, has previously been identified in relation to Crohn’s disease, (28) and there has been evidence using mouse models that *KSR1* kinase activity becomes active at the onset of inflammatory bowel disease. (60) Blood monocytes have been found to be important in innate immunity and inflammatory processes, and changes to the composition of monocytes in the blood have been associated with Crohn’s disease. (40) *PRKCQ*, which provided evidence of a genetically predicted effect on asthma risk, is a member for the protein kinase C family and encodes the enzyme protein kinase C theta which has an important role in the regulation of signalling pathways and the activation of T cells. Moreover, *PRKCQ* has been identified as having a crucial role in autoimmunity through T cell activation. (61, 62) Findings in this study provided evidence of genetic colocalization for this gene with asthma in T cells but none of the other immune cell types assessed, suggesting that *PRKCQ*’s role in conferring autoimmune disease risk may be confined to T cells. (39) *C2orf43* was identified in relation to prostate cancer and provided evidence of colocalization for prostate cancer in monocytes and T cells. It has previously been shown that loss of *C2orf43* may be associated with risk of prostate cancer, and is a gene found to be expressed in lysates of human monocytes and monocyte-derived macrophages. (42)

We found that the 538 effects which provided evidence of genetic colocalization using cell-type specific gene expression were enriched for certain disease outcomes. In particular, various cells types were enriched amongst rheumatoid arthritis loci and in particular for activated naïve CD8^+^ T cells. Increased levels of these cells have been previously observed in the peripheral blood of patients with rheumatoid arthritis. (43) It has also been suggested that CD8 T cells have a role in the initiation and maintenance of rheumatoid arthritis. (63) Monocytes were enriched amongst multiple sclerosis loci, which have previously been implicated in the pathology of this disease by increasing levels of cytokines leading to increased cellular activation and proliferation, tissue damage and altered blood brain barrier. (44) Regulatory memory T cells were enriched for breast cancer which have been identified as markers of poor prognosis of breast cancer by accumulating the tumour tissue and peripheral blood leading to suppression of the immune system. (45)

Our PheWAS analysis highlighted genes which are involved in conferring risk of multiple autoimmune disease outcomes, such as well-established autoimmune locus *IL24*. This gene is in the interleukin family of cytokines which are involved in signalling in the immune system and regulating immune cells,(30) *IL24* has been identified as a key mediator for both pro-inflammatory diseases and allergic disorders. (64) Similarly, *CASP10* was identified in relation to various cancer outcomes in this analysis supporting previous findings. (33) *CASP10* encodes for enzyme caspase-10 which is a member of the caspase family which have a role in cell apoptosis, (65) and have been considered as potential therapeutic targets for cancer. (66)

There were also loci that had specific genetic effects on certain outcomes across the disease spectrum. For example, *PRKCQ* provided robust evidence of an effect on asthma and allergy outcomes, and as previously mentioned this gene has been shown as important in asthma pathology. We also note that our PheWAS results may help flag pleiotropic loci which should be valuable for therapeutic validation endeavours. As an example, we demonstrate that previously postulated target *TPM3* for cancer therapy had genetically predicted effects on various disease endpoints, some of which had the opposite direction of effect to lower cancer risk. Evidence of horizontal pleiotropy may be useful in terms of deprioritising drug targets, whereas those which appear to be more specifically associated with disease, such as *PRKCQ*, may be worthwhile prioritising and pursuing further. However, results from our genetic analysis are but one line of evidence to be used in conjunction with findings from other studies such as functional wet lab work.

Although there various strengths to our study there are also limitations. Firstly, the immune-cell specific gene expression datasets are modest in scale compared to GWAS and thus do not explain a large proportion of heritability. (67) Therefore, the genetic effects that are not supported with colocalization evidence may be due limited by low power for our immune-cell datasets. Future datasets generated at scale once technologies become more feasible should facilitate more comprehensive evaluations of cell-type specific regulatory mechanisms. Moreover, larger sample sizes from cell-type specific datasets would allow a higher degree of confidence that signals identified using whole blood are not subject to molecular pleiotropy (i.e. neighbouring genes being co-expression making pinpoint the causal gene at a locus challenging). Another limitation is horizontal pleiotropy which may play a role in these results despite the use of colocalization, where a causal variant influencing immune-cell expression and disease trait acts via two separate biological pathways. Finally, gene expression was not derived from disease related datasets and were mostly from “healthy” individuals. As such future work using genetic effects on gene expression derived from individuals diagnosed with autoimmune disease or cancer may be potentially capture signatures not detected by our anaylses.

The results of this study provide a map of genetically predicted regulatory mechanisms that influence disease outcomes with an immune basis. These findings should prove valuable for future studies to further characterize susceptibility loci and translate genetic evidence for disease prevention and treatment purposes.

## Methods and materials

### Data resources

GWAS summary statistics were obtained from the MRC IEU analyses of UK Biobank data and consortia who have made their results publicly available (a full list can be found in **Supplementary Table 1**). We obtained eQTL data derived 31,684 blood and peripheral blood mononuclear cell samples from 37 datasets made available by the eQTLGen consortium (http://www.eqtlgen.org), which was accessed on 18/10/2018. (68) The majority of the samples came from individuals with a European ancestry and results were meta-analyzed using a weighted Z-score method. Immune cell-specific eQTL data was obtained from the BLUEPRINT and DICE project, (13, 14) 3 cell types from BLUEPRINT and 15 from DICE (a full tables of cell types can be found in **Supplementary Table 15**).

### Statistical analysis

We performed two-sample MR using the inverse-variance weighted (IVW) MR method transcriptome-wide whilst accounting for linkage disequilibrium structure to investigate the relationship between gene expression, using weakly correlated genetic cis-instruments (16), and 16 disease outcomes. The IVW MR method follows three assumptions; that the genetic instruments are associated with the exposure, that the instrument is not pleiotropic and doesn’t have an effect on the outcome through a pathway other than via the exposure, and that the instruments are no associated with confounders. (69) A Bonferroni corrected threshold was used to identified gene expression with strong evidence for a relationship with an outcome, this threshold was calculated using the number of genes included in the MR analysis for each outcome (Bonferroni corrected threshold=0.05/number of genes).

We then performed a leave-one-out MR analysis for effects which survived Bonferroni correction, which involved reapplying the IVW method after removing each SNP in turn with replacement, to determine whether any individual SNPs were driving genetically predicted effects. If the number of SNPs was 2 or less the Wald ratio MR method was applied instead. Genetic colocalization was undertaken using the ‘coloc’ R package using default parameters. This was to investigate whether the causal variant at a locus responsible for conferring disease risk was also driving variation in gene expression derived from cell-type specific datasets. In total, we applied the coloc method at each locus identified by MR analyses systematically using gene expression data derived from 18 types of immune cells from the BLUEPRINT and DICE consortia. A threshold for PPA scores of ≥80% was used to indicate evidence of a shared a common causal variant between disease outcome and cell-type specific gene expression.

For all effects which provided evidence of genetic colocalization in the previous analyses, we used a hypergeometric test to assess the enrichment of cell-type specific gene expression for each disease outcome. The “phyper” R package was used to perform enrichment analysis. Background comparisons were based on the other loci identified by our MR analyses which did not provide evidence of colocalization with gene expression from the same cell-type. For autoimmune disease outcomes, we used loci for the other autoimmune disease outcomes in the background set, whereas for cancer outcomes we used loci for the other cancer outcomes. (70) Lastly, we performed a phenome-wide association study (PheWAS) using two-sample MR analysis with the same analysis pipeline applied in the initial MR analysis which accounted for local LD structure. However, this analysis assessed the relationship between the genes that were detected in the initial MR analysis and 320 health outcomes which were curated from a previous set of 700 outcomes to ease multiple testing burdens. (71) A list of the 320 outcomes can been found in **Supplementary Table 10**.

All analyses were undertaken using R version 3.6.1 and 3.6.2. MR analyses were conducted using the ‘TwoSampleMR’ and ‘MendelianRandomization’ R packages. (27, 72) Plots illustrating Manhattan and PheWAS plots were generated using ggplot2, (73) locus zoom plots gassocplot (https://github.com/jrs95/gassocplot) and enrichment plots using pheatmap (https://cran.r-project.org/web/packages/pheatmap/index.html). The web application to disseminate findings was constructed using the ‘shiny’ R package.

## Data Availability

All the results from this study are available in our web application (http://mrcieu.mrsoftware.org/immuno_MR/), and code for analyses are available from the 'TwoSampleMR', 'MendelianRandomization' and 'coloc' R packages. Summary statistics for GWAS traits were obtained from publicly available resources as described in Supplementary Table 1 and also from the OpenGWAS platform (https://gwas.mrcieu.ac.uk/). Data on gene expression data from whole blood was downloaded from https://www.eqtlgen.org/ and cell-type specific gene expression was obtained from https://dice-database.org/ and http://dcc.blueprint-epigenome.eu/). The reference panel used in this study was created using data from the UK Biobank (app 15825).

https://gwas.mrcieu.ac.uk/

https://www.eqtlgen.org/

https://dice-database.org/

https://dice-database.org/

## Acknowledgements

We are extremely grateful to the eQTLGen, BLUEPRINT, DICE and GWAS consortia who kindly made their data available for the benefit on this study. CP is supported by a Wellcome Trust PhD studentship in Molecular, Genetic and Lifecourse Epidemiology [108902/B/15/Z].TGR is a UKRI Innovation Research Fellow (MR/S003886/1). This work was supported by the MRC Integrative Epidemiology Unit which receives funding from the UK Medical Research Council and the University of Bristol (MC_UU_00011/1). The reference pnale used in this study was created using data from the UK Biobank (app 15825).

## Materials and Correspondence

This publication is the work of the authors and TGR will serve as guarantor for the contents of this paper.

## Supplementary Tables

Table S1 Table of GWASs for outcomes investigated

Table S2 List of genes and number of cis instruments investigated

Table S3 Table of Bonferroni corrected thresholds

Table S4 MR results that passed the bonferroni corrected threshold

Table S5 MR leave-one-out analysis

Table S6 Colocalisation results

Table S7 Colocalisation results for genes in Figure 3

Table S8 Enrichment results for autoimmune outcomes

Table S9 Enrichment results for cancer outcomes

Table S10 PheWAS health outcomes

Table S11 PheWAS results for PRKCQ

Table S12 PheWAS results for IL24

Table S13 PheWAS results for CASP10

Table S14 PheWAS results for TPM3

Table S15 Table of BLUEPRINT and DICE cell types

